# Rationale and prognosis of repurposed drugs with risk stratification of COVID-19 patients requiring Oxygen supplementation: A systematic review and meta-analysis

**DOI:** 10.1101/2020.10.04.20206516

**Authors:** Esther Jebarani Elangovan, Vanitha Shyamili Kumar, Adhithyan Kathiravan, Raghav Mallampalli, Tiju Thomas, Gnanasambandam Subramaniyam

## Abstract

**Background:** The rising number of trials on repurposed dugs in COVID-19 has led to duplication and a need for curation of available outcomes from treatments that have been followed across the world. We have conducted a systematic review and meta-analysis that focus on evaluating the clinical outcomes of repurposed interventions against COVID-19.

**Methods:** Random effects model was adopted to estimate overall treatment effect and heterogeneity. Meta-regression was performed to study the correlation between comorbid conditions and non-invasive or invasive ventilation requirement.

**Results:** Twenty-nine articles met our eligibility criteria. In subgroup analysis, Tocilizumab was highly significant with lower mortality rate (OR 27.50; 95%CI [5.39-140.24]) of severe COVID-19 patients. Hydroxychloroquine and Lopinavir-ritonavir was found to be inefficacious in severe patients (OR 0.64; 95%CI [0.47-0.86] and 1.40 [0.71-2.76]). Dexamethasone had marginal effect on overall mortality rate (OR 1.19; 95%CI [1.05-1.35]). The meta-regression shows a positive correlation between prevalence of patients on Tocilizumab in non invasive support and hypertension condition (P = 0.02), whereas a negative correlation was identified with patients having lung disease (P = 0.03).

**Conclusion:** Overall, our study confirmed that tocilizumab may probably reduce the mortality rate (<10%) of severe COVID-19 patients than other interventions. Further, reduce the risk of requiring non-invasive ventilator support in patients with comorbid condition of lung disease. Hydroxychloroquine and Lopinavir-ritonavir has no clinical benefits in severe COVID-19. A high quality evidence is required to evaluate the usage of Serpin + Favipiravir combination in severe or critical COVID-19.

## 1. Introduction

The first incidence of novel coronavirus (COVID-19) was identified in patients with severe respiratory disease in Wuhan, China. Since then, COVID-19 outbreak has grown to 32.7 million cases resulting in 991,224 deaths as on 27th September, 2020 across the world. The effect of and response to the virus is varied based on the immune systems [1] environment risks [2], pre-existing health conditions [3, 4], sex differences [5] and so on across different populations and different countries. The virus has debilitated the global community and will continue to do so until an effective vaccine or antiviral is developed.

The ability of the virus to spread rapidly, causing increased risk of deaths in patients with existing health conditions has been considered to be the most alarming feature of COVID-[6,7] At present, there are no targeted therapies or vaccines available for COVID-19. Based on previous outbreaks like Severe Acute Respiratory Syndrome Coronavirus (SARS-CoV) in 2003 [8–10] and the Middle East Respiratory Syndrome Coronavirus (MERS-CoV) in 2012 [11–13], medical community has been working swiftly by repurposing the drugs and evaluating their efficacy in the treatment of the novel coronavirus. Current treatments are primarily based on factors such as combination of drugs, disease severity and respiratory support. Treatment paradigms follow WHO guidelines or those offered by health authorities in each country. Drugs such as Hydroxycholoroquine (HCQ) which were initially reported to be effective in treatment were later found to have limited effects [14]. Later analyses have shown other immune therapies, repurposed antivirals have positive effects on patients in particular phases of treatment [15,16].

Lack of specific treatment and drug therapies for COVID-19, has led the scientific and medical communities to run several drug trials in the past seven months. These studies generated a huge collection of data regarding the drug efficacy, adverse effects and its specificity towards certain populations. The aim of this study is to design and implement a data driven meta-analysis of existing literature and available outcomes regarding treatment for novel coronavirus. We have explored the effect of emerging treatments widely followed by present medical guidelines across countries including drugs like Tocilizumab (TCZ), Remdesivir (RM), Favipiravir (FPV), Dexamethasone (DM), Lopinavir-ritonavir (LPV-r) and Convalescent Plasma (CP) therapy on patients at different severity levels of COVID-19. In addition, we have attempted to manifest the correlation of drug’s ability to treat patients on invasive and non-invasive oxygen support with comorbid conditions like hypertension, diabetes, and cardiovascular diseases. We believe that our findings may help the medical and scientific community to better understand the association of temporal relationships of drug usage during different stages of disease on patients with pre-existing health conditions.

## 2. Materials and Methods

We report a systematic review and meta-analysis, as per the recommendations of PRISMA (Preferred Reporting Items for Systematic Reviews and Meta-Analyses) statements [17]. Our work has not been registered under PROSPERO.

### 2.1 Search Strategy and Study Selection

We systematically searched databases including PubMed, medRxiv and Scopus for research articles published from anytime up to 1st August, 2020. An advanced search was performed with the following keywords: (Remdesivir OR Favipiravir OR Lopinavir/Ritonavir OR Tocilizumab OR Dexamethasone OR Convalescent Plasma OR Hydroxychloroquine OR Arbidol OR Corticosteroids) AND (“COVID-19” OR “SARS-CoV-2” OR “2019 nCoV”) AND (Moderate OR Severe OR Critically ill OR Hospitalized OR Oxygen Therapy OR Invasive Mechanical Ventilation). The drug names in the keywords were selected from the World Health Organization (https://www.who.int/), Indian Council of Medical Research (https://www.icmr.gov.in/), and Ministry of Health and Family Welfare (https://www.mohfw.gov.in/) COVID-19 protocol guidelines. Additionally, we screened the references of the included articles to obtain more relevant papers for the study.

The following articles were excluded from the analysis: duplicates, review papers, editorials, letters, comments, other language manuscripts and studies tested on in-vitro cell culture and in-vivo animals. Inclusion criteria for the study were: i) randomized (RCT) or non-randomized clinical trials (nRCT), prospective or retrospective observational studies (cohort study and case series) ii) research articles, preprints and preliminary reports with comparators (Treatment *Vs*. Control) or combination of treatment modalities or studies without control group iii) study population could be any age, sex and any region in the world, diagnosed with COVID-19 with either laboratory test-confirmed or Chest computer tomography (CT) iv) any one of these outcomes reported: mortality rate, recovery rate, viral clearance period, clinical improvement of patients in oxygen therapy or invasive mechanical ventilation (IMV) after drug treatment.

### 2.2 Data extraction

Data extracted from the included study articles were updated on a google spreadsheet. Any ambiguity in data extraction was clarified by discussion and consensus of the authors. Clinical insights were consistently sought. The following features were extracted: author, study type, date of publication, study period, study place, drug name, cohort size, gender, age, severity condition (mild, moderate, severe and critical); treatment combination, time from symptom onset to the treatment, dosage details; precondition of patients (PaO2:FiO2 and SpO2 levels), comorbidity, patients requiring respiratory support such as low flow oxygen support, high flow or Non-Invasive mechanical Ventilation (NIV), IMV or extracorporeal membrane oxygenation (ECMO) (during admission and follow-up); clinical improvement length, viral clearance period; mortality rate; recovery rate and adverse effects. All the data were individually extracted for subgroups (treatment group, control or comparator) and overall outcomes of all the treatments were summarized together and plotted using ggplot2 [18] in R.

### 2.3 Risk of bias assessment

We used RoB 2.0 [19] and ROBINS-I [20] tools of Cochrane risk of bias assessment for evaluating RCTs and observational or nRCTs. Robvis package [21] from R was used for the visualization of risk bias assessment. The Newcastle-Ottawa scale (NOS) was used to assess single-arm studies [22]. The use of this scale is more sensible to control the quality level of the cohort study [23].

### 2.4 Statistical analysis

To evaluate the drug treatment effects between control and test groups, an odds ratio was obtained to measure probability of events occurring between groups at different disease severity levels. The intervention effect distribution was estimated using the Random Effects Model (REM), which is provided as an estimate of 95% confidence interval. I^2^ statistic was used to measure heterogeneity within and between studies. This was performed using metafor package V2.4.0 [24] in R V4.0.2. Overall proportion of all single armed studies were calculated using metaprop [25] program in R. Meta-regression was performed to assess the correlation of drug efficacy in patients at different oxygen therapy stages with comorbid conditions using lm function in R. A p-value of ≤ 0.05 is considered as statistically significant in all our studies unless stated.

## 3. Results

Overall, 15,831 records were identified by our searches. On removing redundant entries, 3095 numbers of papers were retained. After exclusion of review articles, 29 clinical studies (24 published and 5 pre-prints) met our eligibility criteria (PRISMA flow chart Figure 1). All these were available online between 11th March, 2020 and 22nd July, 2020. After definitive selection of articles, there were eight therapeutic agents in total found from 29 studies having 14,114 COVID-19 patients involved were compared in our meta-analysis. These eight agents included TCZ (trialed on 684 participants), CP (95), RM (158), DM (2104), HCQ (912), LPV-r (215), Arbidol (137), FPV (2383) and Standard care or placebo (6344). Nine (6 published and 3 pre- prints) of 29 included studies were single-arm trials without a comparator. The description of the participants, treatment, clinical outcomes and the key findings of all the included studies are presented in Table 1.

**Table 1.** Characteristics of Baseline characteristics and key outcomes of included studies

| Author | Study type(Country) | Participants | Treatment | Comparison | Outcomes | Key findings | Overall risk of bias |
| --- | --- | --- | --- | --- | --- | --- | --- |
| **Tocilizumab** |  |  |  |  |  |  |  |
| Campochiaro et al. [27] | Single-centre retrospective observational study (Italy) | Severe COVID-19 (n=65) | Tocilizumab i.v. at 400 mg once plus standard care. Second dose 400 mg after 24 h in case of respiratory worsening (n=32) | Standard care without tocilizumab (n=32) | Survival; 2 point decrease six-category ordinal scale; clinical improvement at day-28; adverse events | At day 28 TCZ *Vs.* STD: No significance in mortality (15% *Vs.* 33%) (p = 0.15), where age $\geq 75$ as predictors of death and clinical improvement (69% *Vs.* 61%)(p = 0.61), where PaO<sub>2</sub>:FiO<sub>2</sub> $\geq 100$ as predictors; Bacteremia (13% *Vs.* 12%) | High risk (ROBINS -I) |
| Kewan et al. [66] | Single-centre retrospective observational study (USA) | Severe COVID-19 (n=51) | Tocilizumab i.v. 8 mg/kg and received a maximum of 400 mg for 60 min once plus standard care (n=28) | Standard care without Tocilizumab (n=23) | Clinical improvement; 2 point decrease six-point scale or live discharge; adverse events | TCZ *Vs.* STD: Shorter duration of IMV (7 *Vs.* 10 days) and vasopressor (2 vs 5 days) in TCZ group; age $\geq 65$ years on IMV had higher clinical improvement rate (40% *Vs.* 13%, p = 0.20); No significance in clinical improvement 76.5% *Vs.* 79.4% | High risk (ROBINS -I) |
|  |  |  |  |  |  | by day 21 |  |
| Guaraldi et al. [41] | Single-center retrospective observational cohort study (Italy) | Severe COVID-19 (n=544) | Tocilizumab i.v. 8 mg/kg twice 12h apart (n=179) | Standard care without tocilizumab (n=365) | Risk of death or invasive mechanical ventilation, adverse events or severe adverse events | TCZ *Vs.* STD: Significant reduction in risk of IMV or death in TCZ group (adjusted hazard ratio 0·61, 95% CI 0·40–0·92; p=0·020); mortality reduced in TCZ treatment 13 (7%) *Vs.* 73 (20%) (p<0·0001); new infections (13% *Vs.* 4%, (p<0·0001)) | Moderate risk (ROBINS -I) |
| Klopfenstein et al. [42] | Single-center retrospective case-control study (France) | Critically ill COVID-19 (n=45) | Tocilizumab 1 or 2 doses plus standard treatment (n=20) | Standard treatment without tocilizumab (n=25) | ICU admissions; death; IMV; discharge | Reduction in ICU admission/death (25% *Vs.* 72%, p = 0·002) or requiring IMV (0% *Vs.* 32%, P = 0·006) in TCZ group than control | Moderate risk (ROBINS -I) |
| Rossi et al. [40] | Multi-centre retrospective observational study (Italy) | COVID-19 patients with early stage respiratory failure (n=158) | Tocilizumab i.v. 400 mg or s.c.324 mg plus standard therapy (n=90) | Standard therapy without tocilizumab (n=68) | Clinical and laboratory results at day 5; IMV; death | Reduction of death (94%) in TCZ group at low dose in early stage respiratory failure; no difference in two administration modes (p = 0·292); no adverse events | Moderate risk (ROBINS -I) |
| Capraa et al. [26] | Single-center retrospective observational study (Italy) | COVID-19 patients requiring respiratory support except IMV (n=85) | Tocilizumab i.v. 400 mg once or s.c. 324 mg once plus standard care (n=62) | Standard care without Tocilizumab (n=23) | Survival; respiratory followup; infections | TCZ Vs. STD: greater survival rate (HR 0.035; 95% CI, 0.004 to 0.347; p = 0.004), recovery (92% and 42.1%) and still hospitalized recovery (64.8% Vs. 0%) in TCZ group; no infections | Moderate risk (ROBINS -I) |
| Tomasiewicz et al. [43] | Multi-centre retrospective observational study (Poland) | Severe COVID-19 (n=28) | Tocilizumab i.v. 800 mg once (n=28) | No comparison | Clinical status at 24 hrs, 1 and 2 weeks; laboratory data; adverse events | TCZ increases SpO2 levels from 89% (Day 1) to 97% (day 10) (p ≤ 0.001); 2 weaned mechanical ventilation within 24 hrs after first dose; 84% chest CT improvement in 2 weeks; death or slow improvement in patients associated with 2 or more comorbidities | Poor quality (NOS) |
| Sciascia et al. [44] | Multi-centre prospective pilot study (Italy) | Severe COVID-19 adults (n=63) | Tocilizumab i.v. 8 mg/kg or s.c. 324 mg. Second administration s.c. | No comparison | Laboratory data; IMV; Safety | TCZ administration significantly increased survival | Poor quality (NOS) |
|  |  |  | 162 mg before 24 h (n=63) |  |  | within 6 days (HR 2.2 95%CI 1.3-6.7, p<0.05) and PaO2:FiO2 ratio from 152±53 (day 0) to 302.2 ±126 (day 14) (p<0.05); improvement in D-dimer, ferritin, C-reactive protein levels and lymphocytes count (p<0.05) |  |
| Toniati et al. [34] | Single-center prospective case series (Italy) | Severe COVID-19 (n=100) | Tocilizumab i.v. 8mg/kg twice 12 hr apart (n=100) | No comparison | Laboratory data at day 10; ICU improvement; severe adverse events | TCZ administration showed 58% rapid improvement at 24-72hr in respiratory severity condition than pre-TCZ; By day10: 77% improved respiratory condition, 54% suspended non-invasive ventilation, 74% of 43 ICU patients improved; 20% mortality; 3% Severe adverse effects | Poor quality (NOS) |
| Montalva et al. [35] | Single-center prospective cohort study (Spain) | Hospitalized adults with severe COVID-19 | Tocilizumab 600 mg (>75Kg) or 400 mg (<75Kg) once | No comparison | Death at Day7; admission to Intensive Care Unit (ICU), acute Respiratory Distress Syndrome (ARDS) and respiratory insufficiency ; adverse events | TCZ: Mortality(26.8%) was more frequent in test group with ARDS (HR 3.3; 95% CI, 1.3 to 8.5), respiratory failure was present (HR 3.13; 95% CI, 1.3 to 7.8); no serious adverse events | Poor quality (NOS) |
| **Convalescent plasma** |  |  |  |  |  |  |  |
| Li et al. [29] | Multi-centre randomized open label study (China) | Severe or life-threatening COVID-19 (n=103) | Convalescent plasma transfusion 10 mL first 15 min, then upto 100 mL/ hr based on the patient's risk or tolerance, plus standard treatment (n=52) | Standard treatment without plasma (n=51) | Discharge; 2 points less 6-point disease severity scale; viral PCR test at 24,48 and 72 hrs and post-hoc day 7, 14 and 28 | Significant increase in viral PCR negative conversion rate at 72 hrs in CP group than STD (odds ratio (OR) 11.39 (95% CI, 3.91-33.18) P < .001); no significance in 28-day mortality (p= .30) or discharge (p= .12); adverse events in patients after transfusion were cured | High risk (RoB 2.0) |
| Gharbharan et al. [28] | Multi-centre randomized open label study | COVID-19 positive (RT-PCR test) (n=86) | Convalescent plasma i.v. 300ml plus standard care (n=43) | Standard care without plasma (n=43) | Overall mortality upto 60 days; 8-point | Recepients had high antibody titers than donors before transfusion; | High risk (RoB 2.0) |
|  | (Netherlands) |  |  |  | disease-severity scale at day 15; safety outcomes | no serious adverse events; clinical improvement (p=0.58) or mortality (p=0.95) were not significant in CP group and STD; early symptom onset greatly benefited |  |
| **Remdesivir** |  |  |  |  |  |  |  |
| Goldman et al. [50] | Multi-centre randomized open label study (United States, Italy, Spain, Germany, Hong Kong, Singapore, South Korea, and Taiwan) | Severe COVID-19 (n=397) | Remdesivir i.v. 200 mg once on day 1 ,100 mg once for subsequent 4 days (n=200) | Remdesivir i.v. 200mg once on day1 ,100mg once for subsequent 9 days (n=197) | Clinical status at day 14 on 7-point ordinal scale; Duration of clinical improvement; Clinical recovery and mortality rate; adverse events and serious adverse events | Remdesivir 5-day *Vs.* 10-day: No significance in clinical improvement (64% *Vs.* 55%) ;no difference in IMV patients; men has worse outcomes(68%) | High risk (RoB 2.0) |
| Grein et al. [49] | Multi-centre randomized open label study (United states,canda,japan) | Severe COVID-19 (n=53) | Remdesivir i.v. 200 mg once on day 1 ,100 mg once on day 2 to day 10 (n=53) | No comparison | Cumulative incidence of clinical improvement till day 36; mortality; adverse events or serious adverse events | By Day 28: 68% clinical improvement in oxygen support patients; high mortality in age $\geq 70$ | Fair quality (NOS) |
| Beigel et al. [30] | Multi-centre randomized double-blind study (USA, Denmark, UK, Greece, Germany, Korea, Mexico, Spain, Japan, Singapore) | Hospitalized adults with lower respiratory tract evident (n=1059) | Remdesivir i.v. 200 mg once day1 followed by 100 mg day 2-10 once (n=538) | Placebo (n=521) | Clinical recovery; 8-category ordinal scale; adverse or serious adverse events | Shorter recovery duration in remdesivir treated than placebo (median 11 Vs. 15 days)(P<0.001); by day 15: improvement were higher in remdesivir group (rate ratio(RR) 1.32; 95% CI, 1.12 to 1.55, P<0.001); Ordinal score 5 (low oxygen support) benefited the most (RR 1.47 (95% CI, 1.17 to 1.84); age 18 to <40 had highest recovery (RR 2.03 95% CI 1.31- 3.15) | Low risk (RoB 2.0) |
| Wang et al. [31] | Multi-centre randomized double-blind study (China) | Severe COVID-19 adults (n=236) | Remdesivir i.v. 200 mg once day1 followed by 100 mg day2-10 once (n=158) | Placebo (n=78) | Live discharge; all-cause mortality at day-28; clinical improvement duration and a 2 point decrease 6-point scale at day 7, 14 and 28; virological measures; adverse or serious adverse events | No significance in clinical improvement duration (median 21 Vs. 23 days), 28-day mortality (14% Vs. 13%) and negative viral RNA proportions in remdesivir group from placebo | Low risk (RoB 2.0) |
| **Dexamethasone** |  |  |  |  |  |  |  |
| Horby et al. [32] | Multi-centre randomized open label study (UK, USA and Italy) | Hospitalized with Covid-19 (n=6425) | Dexamethasone i.v. or orally 6mg once daily for 10 days plus usual care (n=2104) | Usual care without dexamethasone (n=4321) | 28-day mortality; composite outcome of IMV or death; discharge | IMV deaths lower in dexamethasone group than usual care (29.3% *Vs.* 41.4%); no significance in patients receiving oxygen only (23.3% *Vs.* 26.2%), without oxygen (17.8% *Vs.* 14.0%) and overall mortality (22.9 % *Vs.* 25.7%) at day 28 | Low risk (RoB 2.0) |
| **Hydroxychloroquine** |  |  |  |  |  |  |  |
| Chen et al. [52] | Single-center randomised double-blind study (China) | Mild COVID-19 adults (n=62) | Hydroxychloroquine 200mg twice/day for 5 days plus standard treatment (n=31) | Standard treatment without hydroxychloroquine (n=31) | Time to clinical recovery (TTCR); pulmonary recovery; adverse events | Shorter recovery time of body temperature (2.2 *Vs.* 3.2 days) and cough and significant improvement of pneumonia | High risk (RoB 2.0) |
|  |  |  |  |  |  | (80.6% *Vs.* 54.8%)<br>in<br>hydroxychloroqui<br>ne group than<br>control; no severe<br>side effects |  |
| Tang et al. [53] | Multi-center<br>open label<br>controlled<br>randomised<br>study (China) | Hospitalized<br>Patients with<br>Covid-19<br>(n=150) | Hydroxychloroqui<br>ne 1200 mg daily<br>on day1-3 and<br>800 mg daily for 2-3 weeks plus<br>standard care<br>(n=70) | Standard care<br>without<br>Hydroxychloroqui<br>ne (n=80) | Negative<br>conversion;<br>adverse<br>events or<br>severe<br>adverse<br>events | HCQ+STD *Vs.*<br>STD: No<br>significant<br>difference in<br>probability of<br>negative<br>conversion (85.4%<br>*Vs.* 81.3%); more<br>adverse events in<br>test group | Some<br>concerns<br>(RoB 2.0) |
| Geleris et al. [33] | Single-center<br>observational<br>Study (USA) | Hospitalized<br>patients with<br>Covid-19<br>(n=1376) | Hydroxychloroqui<br>ne 600 mg twice<br>on day 1 and 400<br>mg daily on days2-5 ; Azithromycin<br>500 mg on day1<br>and 250 mg daily<br>on days2-5<br>combination with<br>HCQ (n=811) | Control without<br>hydroxychloroquin<br>e (n=565) | Intubation or<br>death at<br>regular time<br>intervals | HCQ *Vs.* no HCQ:<br>High intubation or<br>death in test group<br>(32.% *Vs.* 14.9%);<br>HCQ use is not<br>associated with<br>high or low risk at<br>intubation or death | Moderate<br>risk<br>(ROBINS<br>-I) |
| **Lopinavir/ritonavir** |  |  |  |  |  |  |  |
| Cao et al. [58] | Single-centre<br>randomized<br>open label<br>study (China) | Hospitalized<br>adults with<br>Severe<br>COVID-19<br>(n=199) | Lopinavir/ritonavir<br>400mg/100mg<br>every 12h plus<br>standard care for<br>14 days (n=99) | Standard care<br>without<br>Lopinavir/ritonavir<br>(n=100) | Clinical<br>improvement<br>duration;<br>Seven-<br>category<br>ordinal scale<br>or discharge;<br>virologic<br>measures; | LPV-r *Vs.* STD:<br>shorter ICU stay<br>(median 6 *Vs.* 11<br>days); no<br>significance in<br>mortality (−5.8<br>95% CI, −17.3 to<br>5.7), clinical<br>improvement (8.8 | Some<br>concerns<br>(RoB 2.0) |
|  |  |  |  |  | safety | 95% CI, -3.3 to 20.9) and viral detectability (60.3% vs. 58.6%) at day 28; 4 gastrointestinal serious adverse events in the LPV-r group |  |
| Li et al. [55] | Single-centre randomized study (China) | Mild/moderate COVID-19 (n=86) | Treatment I: Lopinavir/ritonavir 200/50 mg twice daily for 7-14 days (n=34) OR Treatment II: Arbidol 200mg thrice 7-14 days (n=35) | Standard care without antiviral therapy (n=17) | RT-PCR negative conversion rate or chest CT improvement rate at day 7 and 14; Adverse events | LPV-r Vs. Arbidol Vs. STD: Clinical deterioration (23.5% Vs. 8.6% Vs. 11.8%) at day 7; virological measures similar among all groups (p> 0.05); 12 adverse or one serious adverse events in LPV-r group | High risk (RoB 2.0) |
| **Lpv/r Vs Lpv/r +. Arbidol (test vs control)** |  |  |  |  |  |  |  |
| Deng et al. [56] | Single-center retrospective cohort study (South Korea) | Confirmed COVID-19 without Invasive ventilation (n=33) | Lopinavir/ritonavir 400 mg/100 mg every 12h. (n=16) | Arbidol 200 mg every 8h and Lopinavir/ritonavir orally 400 mg/100 mg twice daily. (n=17) | Clinical improvement outcomes (day14); negative conversion rate of SARS-CoV-2; pneumonia monitoring by chest CT(day7) | LPV-r Vs. LPV-r+ Arbidol: Significance in negative viral load in patients not requiring mechanical ventilations (94% Vs. 53%) and improvement in chest CT (69% Vs. 29%) | High risk (ROBINS-I) |

| Lopinavir/ritonavir vs HCQ |  |  |  |  |  |  |  |
| --- | --- | --- | --- | --- | --- | --- | --- |
| Kim et al. [57] | Single-center retrospective cohort study (South Korea) | Hospitalized patients with COVID-19 (n=65) | Lopinavir/ritonavir 400 mg/100 mg twice daily (n=31) | Hydroxychloroquine 400 mg once daily (n=34) | Time to negative conversion of viral RNA; time to clinical improvement; adverse events or serious adverse events | LPV-r Vs. HCQ: Shorter time to negative conversion of viral RNA (median 21 Vs. 28 days) , no significant in time to clinical improvement (median 18 Vs. 21 days) ,negative conversion is more in patients age <65 | High risk (ROBINS -I) |
| Favipiravir |  |  |  |  |  |  |  |
| Rattanaumpawan et al. [59] | Multi-center retrospective observational study (Thailand) | Hospitalized adult patients with COVID-19 (n=63) | Favipiravir 400 mg twice on day1 followed by 600 mg twice on days2-5 plus standard care (n=63) | No comparison | Clinical improvement; mortality; adverse events | Favipiravir: Rate of clinical improvement is less and slow in patients receiving oxygen (ambient Vs. receiving O2: day7 - 92.6% Vs. 47.2%; day-28 100% Vs. 83.3%), low mortality(7.9%), no severe adverse events | Fair quality (NOS) |
| Doi et al. [58] | Multicenter observational study (Japan) | Confirmed COVID-19 patients (n=2158) | Favipiravir 1600 mg twice on day1 and 600 mg or 800 mg twice daily for the following days(n=2158) | No comparison | Clinical status and outcome at day7, day14 and after 1 month; | Favipiravir: Higher mortality among age group $\geq 60$ (92.37%); less effective for patients receiving | Poor quality (NOS) |
|  |  |  |  |  | adverse events | oxygen |  |
| **Favipiravir Vs. Lopinavir/ritonavir (both with IFN alpha)** |  |  |  |  |  |  |  |
| Cai et al. [67] | Single-centre non-randomized open label study (China) | Confirmed COVID-19 patients (n=80) | Favipiravir 1600 mg twice on Day1 and 600 mg twice daily on days 2–14 (n=45) | Lopinavir 400 mg /Ritonavir 100 mg twice daily (n=35). | Clinical improvement; viral clearance time; chest CT improvement; adverse events | Favipiravir Vs. LPV-r: Shorter time of viral clearance (median 4 Vs. 11 days) ; High significant improvement rate in chest CT (91.4% Vs. 62.22%) ; more adverse events in LPV-r arm; treatment duration in favipiravir arm can be extended if required | Moderate risk (ROBINS-I) |
| **Nafamostat mesylate + Favipiravir** |  |  |  |  |  |  |  |
| Doi et al. [64] | Single-center case series (Japan) | Critically ill COVID-19 (n=11) | Favipiravir i.v. 3600 mg on day 1 and 1600 mg subsequent days plus Nafamostat mesylate continuous i.v. 0.2 mg per kg per hour (n=11) | No comparison | Live discharge; mortality | Favipiravir with Nafamostat mesylate is effective in critically ill patients; low mortality (9%) | Poor quality (NOS) |

| Favipiravir Vs. Arbidol |  |  |  |  |  |  |  |
| --- | --- | --- | --- | --- | --- | --- | --- |
| Chen et al. [60] | Multi-centre prospective randomized controlled open label study (China) | Adult patients with COVID-19 (n=236) | Favipiravir 1600 mg twice on day1 and 600 mg twice daily for the following days(n=116) | Arbidol 200 mg three times daily plus standard care for 7 days (n=120) | Clinical recovery at Day 7; Latency to relief for pyrexia and cough; rate of noninvasive mechanical ventilation; adverse events | Favipiravir(FPV) Vs. LPV-r: No significance in clinical recovery between two groups (P=0.1396), improvement in latency to relief for pyrexia and cough is significant in FPV arm, effective for moderate COVID-19 patients | Low risk (RoB 2.0) |

**Figure 1:**
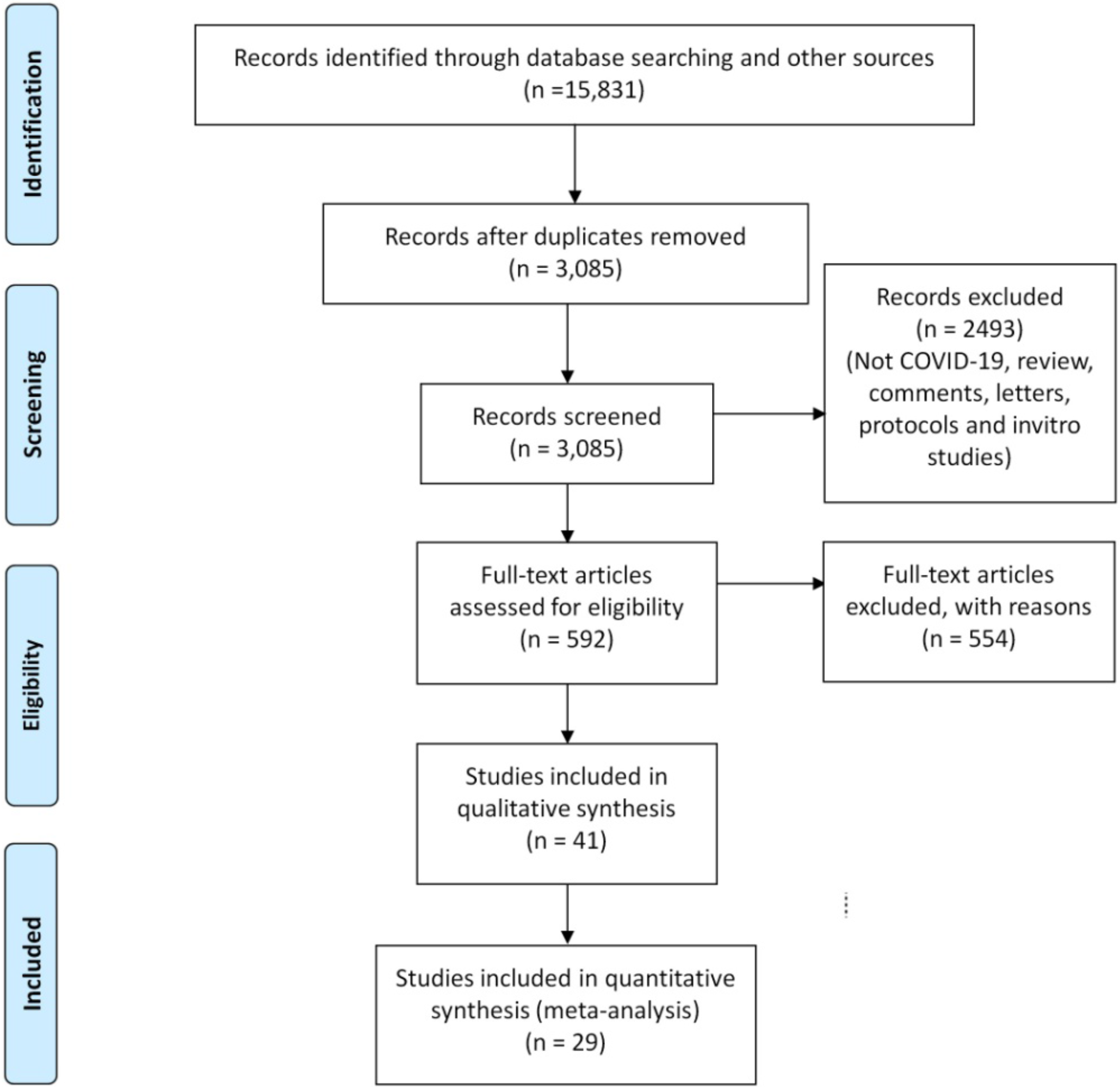
PRISMA Flow diagram.

### 3.1 Risk of bias assessment

The risk of bias score evaluated for 29 included studies comprising 12 RCTs, 1 nRCT, and 16 observational (12 retrospective, 3 prospective and 1 case series). The risks of bias for randomized and non-randomized are presented in Figure 2a and Figure 2b respectively. Risk of bias for single-group studies was assessed using NOS is given in Table S1. The overall judgments for 15 studies were high risk or poor quality, 10 at moderate or fair quality and 4 at low risk of bias (Last column of Table 1).

**Figure 2.**
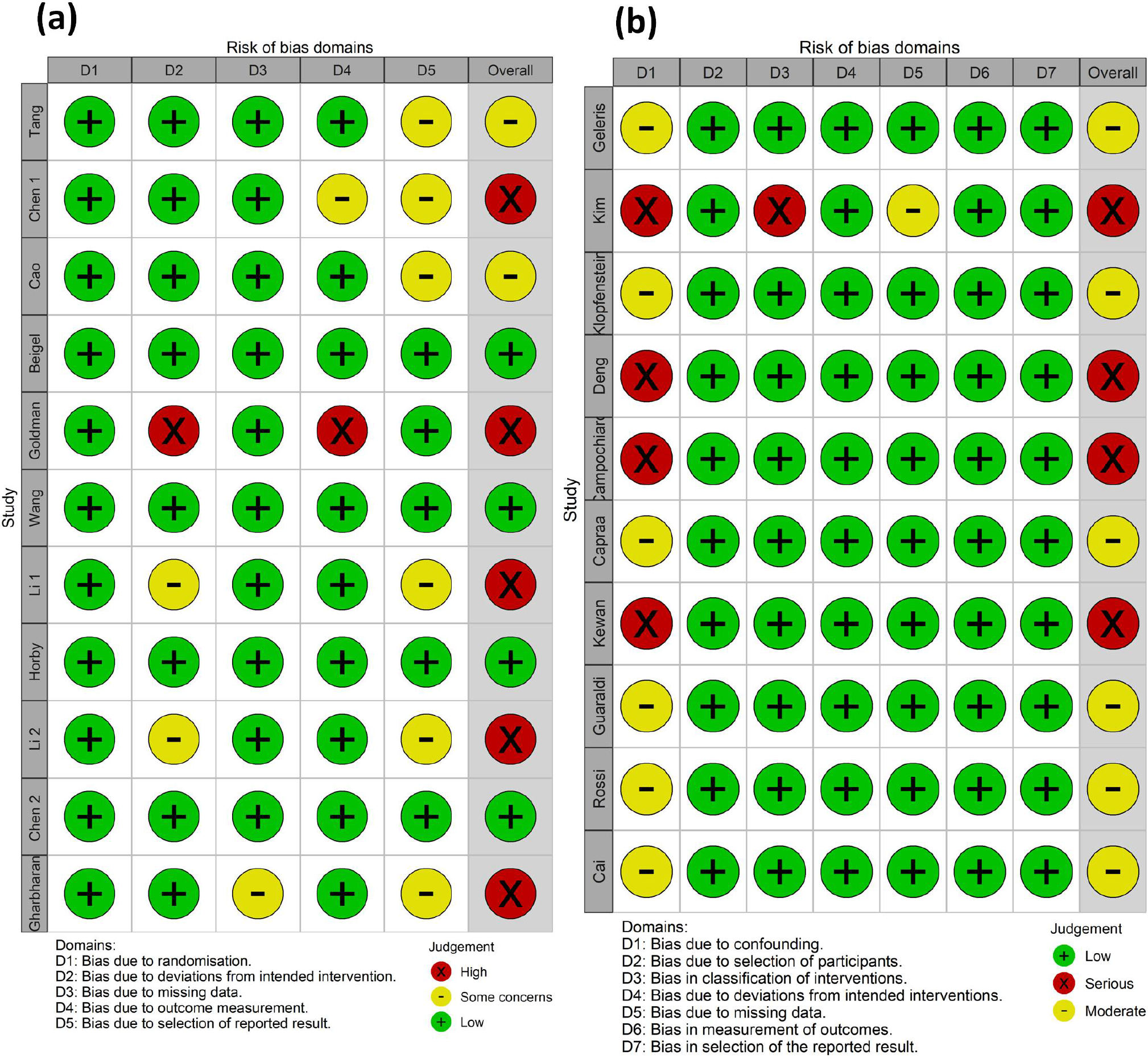
Risk of bias assessment with Cochrane’s collaboration (a) Randomised controlled trails assessment using RoB 2.0 tool (b) Non-randomized controlled trials using ROBINS-I tool

### 3.2 Subgroup analyses based on disease severity levels

By pooling articles based on disease severity levels, studies were subgrouped as 1) mild/moderate 2) severe (Figure 3). Odds ratio of mortality rate for studies including mild/moderate patients found to have higher 95% Confidence Interval (CI), P Value of 1.00 with I^2^ = 0% indicating no observed heterogeneity. However, articles with patients at high risk/severe disease symptoms showed P Value < 0.001 and heterogeneity measure (I^2^) of 87.5%. Most of the studies classified under severe subgroups had small 95% CI and showed positive effects of drugs in the treatment group. On the other hand, a greater part of studies under mild/moderate sub- group indicated the null effect of the drug on the treatment group with long 95% CI.

**Figure 3.**
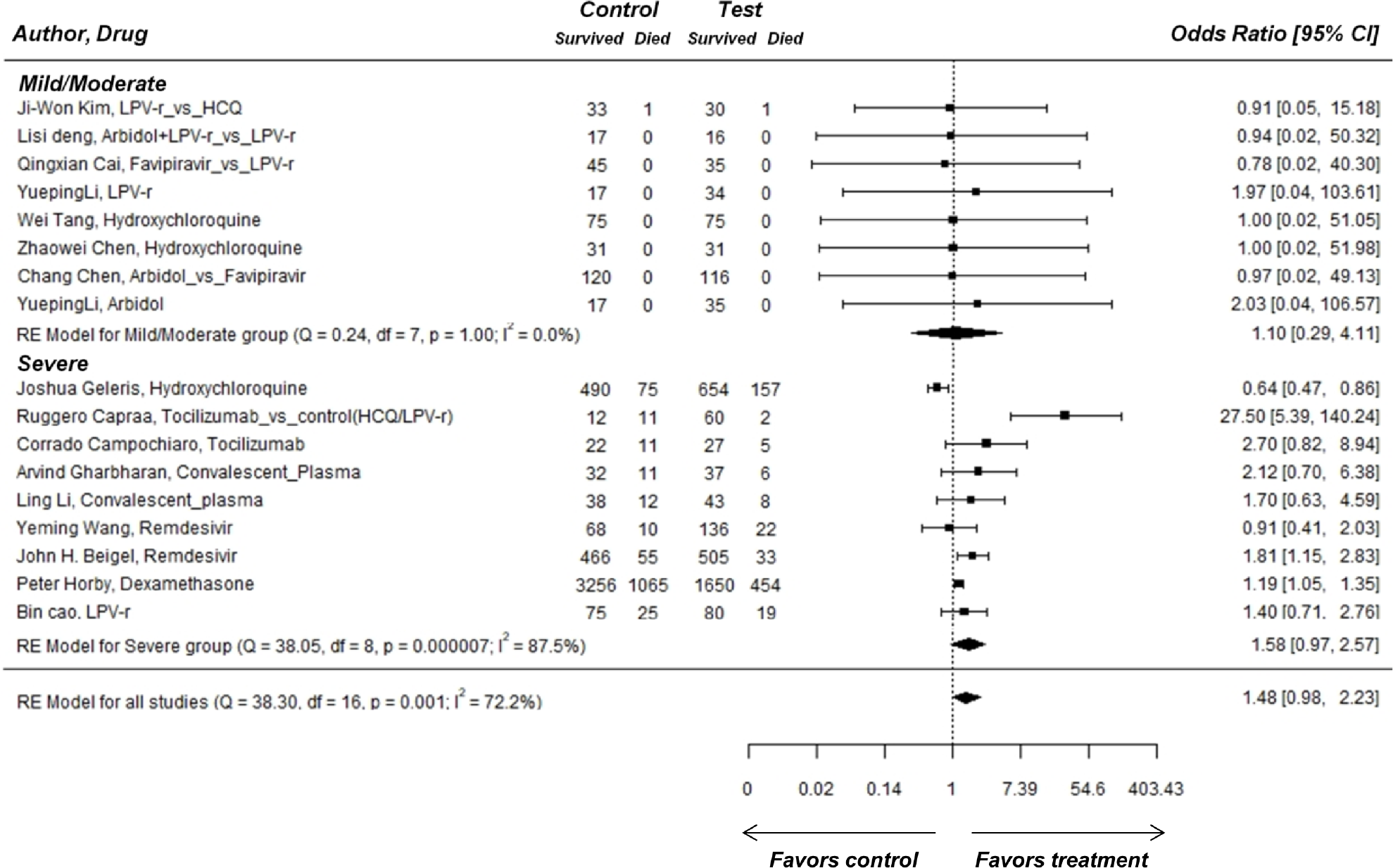
Forest plot comparing mortality of patients between control and test group and subgrouped based on severity level of disease. CI, confidence interval; df, degrees of freedom; I^2^, heterogeneity. Line of no effect is at 1. Each horizontal bar represents upper and lower 95% CI. Squares on the right and left side of the line favor the test and control group respectively. Diamond shows an overall summary of all studies

Of all the drugs included in the severe subgroup, TCZ was found to perform well on patients at high risk (OR 27.50 [5.39-140.24]; 2.70 [0.82-8.94]) [26,27], followed by CP therapy (OR 2.12 [0.70-6.38]; 1.70 [0.63-4.59]) [28,29]. Of two articles published on RM, one showed less response on treatment group [30] (OR 1.81 [1.15-2.83]) while other had null effect (OR 0.91 [0.41-2.03]) [31]. The failure of the latter trial on Remdesivir is due to the small sample size with 2:1 randomisation. Both the studies showed an insignificant mortality rate between control and test groups [30,31]. DM had a marginal positive effect on overall mortality (OR 1.19 [1.05- 1.35]) [32]. HCQ showed negative effects on treatment groups under severe conditions (OR 0.64 [0.47-0.86]). This study [33] on HCQ had a large cohort (n=1376) of hospitalized COVID-19 patients and suggests that HCQ use had no potency for end stage intubation or death. Overall, REM for all studies from all subgroups showed a significant P Value of 0.001 and heterogeneity measure (I^2^) of 72.2%.

### 3.2 Single-group study analysis

Figure 4 shows a visual summary of a patient’s mortality from different drug trial studies with no placebo/control. The results of included single armed studies, with 95% C.I and the pooled proportions are provided. Overall effect of drugs on COVID-19 patients showed a significant mortality proportion of 0.12 (P < 0.01) with heterogeneity measure (I^2^) of 72%. Most of the included studies had lower mortality proportion (< 0.1) except two studies on TCZ [34,35] having > 0.2 proportion. The study with use of FPV treatment shows minimal mortality rate (0.05; 95% CI [0.01-0.13]) [36].

**Figure 4:**
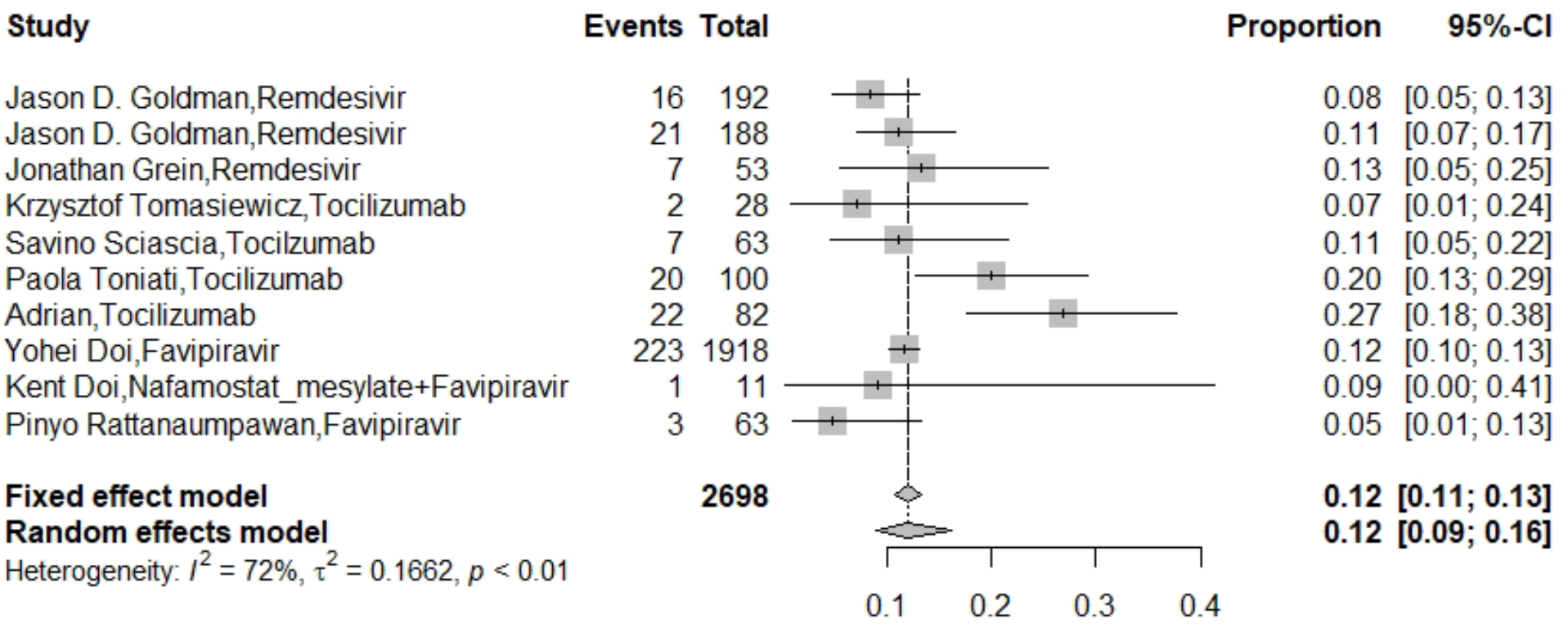
Forest plot of the proportion of death events occurred in different single armed drug trials. Squares on the right side show a higher proportion of death events in the population

### 3.3 Factors associated with the prevalence of patients in different oxygen therapies

We performed meta-regression analysis to understand the association of factors influencing a patient’s dependency on life supporting ventilation treatments. A set of variables that may likely influence the COVID-19 patients landing into non-invasive and invasive oxygen supports were identified. The variables included were time from symptom onset to treatment, treatment duration, mortality rate and comorbidities of patients (hypertension, diabetes, heart disease, lung disease and renal disease). Of all the drugs included, TCZ and RM had enough number of studies (14) with potentially relevant oxygen support information (supplemental oxygen/ NIV/ IMV) to carry out the analysis. Results showing the multivariate meta-regression for all TCZ studies are shown in Figure 5. A total of 376 patients were under respiratory support before the treatment, and 195 remained at the end of treatment. In this multivariate meta-regression model, comorbidities factors showed association with prevalence of patients in non-invasive support (Figure 5a and 5b). There was a significant positive correlation between prevalence of patients on TCZ in non invasive support and hypertension condition (P = 0.02) (Figure 5b). On the other hand, a significant negative correlation with patients having lung disease and their existence in non-invasive support was identified (P = 0.03) (Figure 5b). Additionally, patients with pre-existing diabetes conditions significantly observed to have no association with their prevalence in non-invasive support (P = 0.02) (Figure 5b).

**Figure 5.**
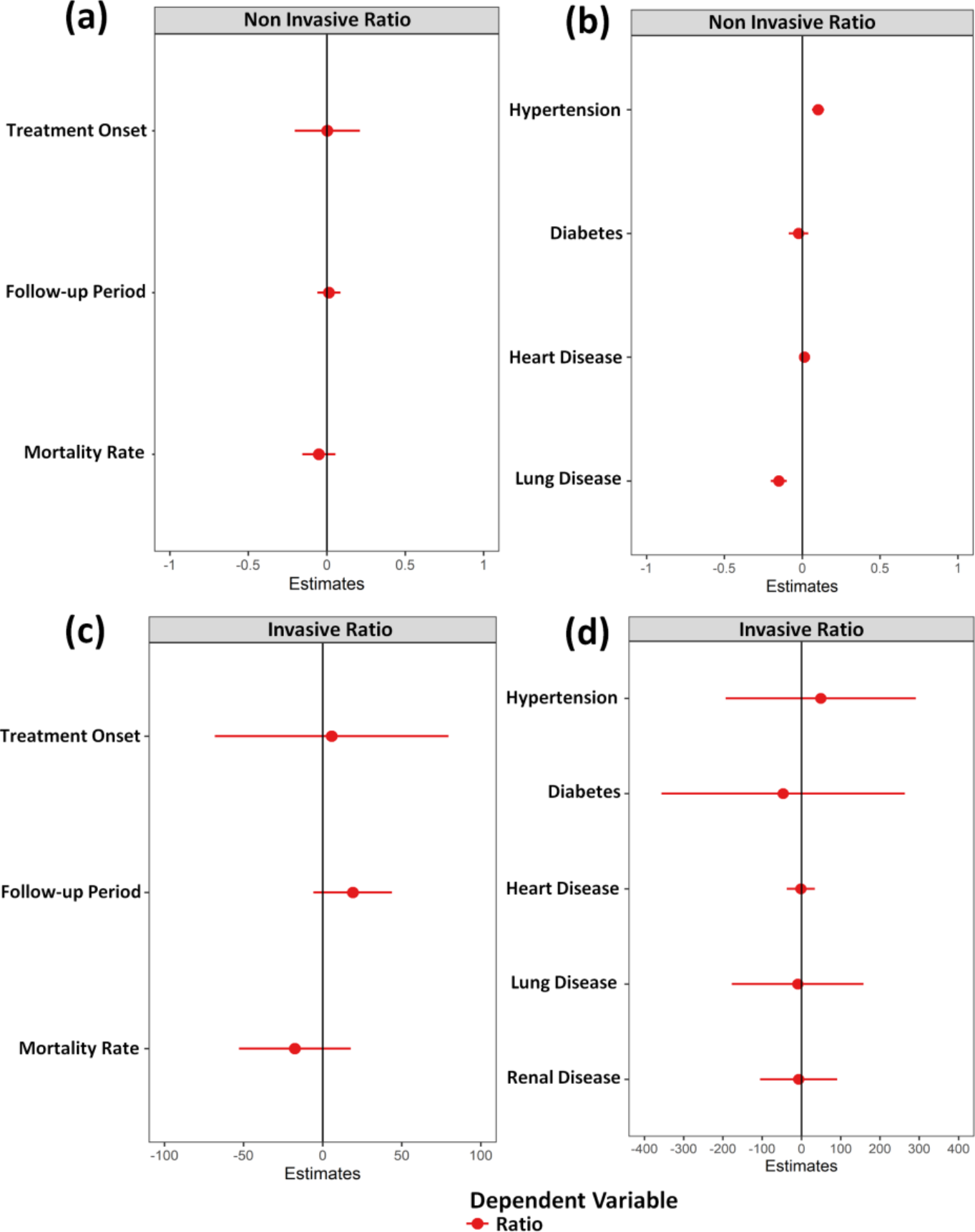
Meta-regression showing the correlation of patient population on TCZ with treatment characteristics and comorbidities. (a) correlation of patients under non-invasive therapy with treatment characteristics (b) correlation of patients under non-invasive therapy with comorbidities (c) correlation of patients under invasive therapy with treatment characteristics (d) correlation of patients under invasive therapy with comorbidities

There was no significant association identified on comparing the patient’s prevalence in invasive ventilation supports (ECMO) and its affecting factors. Similar to the non-invasive case, positive correlation was observed between the number of patients with hypertension and increased risk of falling into invasive oxygen support. On the other hand, patients with diabetes when treated on TCZ showed negative correlation with their prevalence in invasive ventilation support. Furthermore, patients treated with TCZ under invasive support had no association with early treatment onset, and comorbidities conditions including pulmonary, renal and heart diseases. Meta-regression on RM revealed no association of the patient population in oxygen therapy with any of the dependent variables except time from symptoms onset (data not shown).

### 3.4 Combined analysis on comparable clinical outcomes from all studies

The overall clinical outcomes in control and test for each treatment were compared and represented as a bar plot (Figure 6). The percentage of death in the TCZ group was found to be lowest (<10%) among all treatments compared in this study. Additionally, patients in the TCZ group found to have faster recovery (duration of 7days) than the control group and other treatments. Moreover, in the control group many of the patients required invasive support during a follow-up (26%) than at the time of admission (15%), whereas a decrease was observed in the TCZ treatment (18% admission to 17% follow-up). HCQ was the only test group where the death rate relatively scaled up (17%) than in the control group (11%). The clinical outcomes of control and test groups from DM studies showed no significant difference (it was specific for significant reduction of death in IMV support). Although a shorter duration of clinical improvement was observed among TCZ, DM and RM treatments. A good rise in negative conversion rate of the viral RNA (during follow-up) was seen in CP (48% *Vs*. 20%) and FPV (91% *Vs*. 76%) treatments compared to the control group. The studies including RM (20% *Vs*. 27%) and LPV-r (15% *Vs*. 27%) observed to show lower incidence of severe adverse events, the percentage are very much comparable with that of their control group.

**Figure 6.**
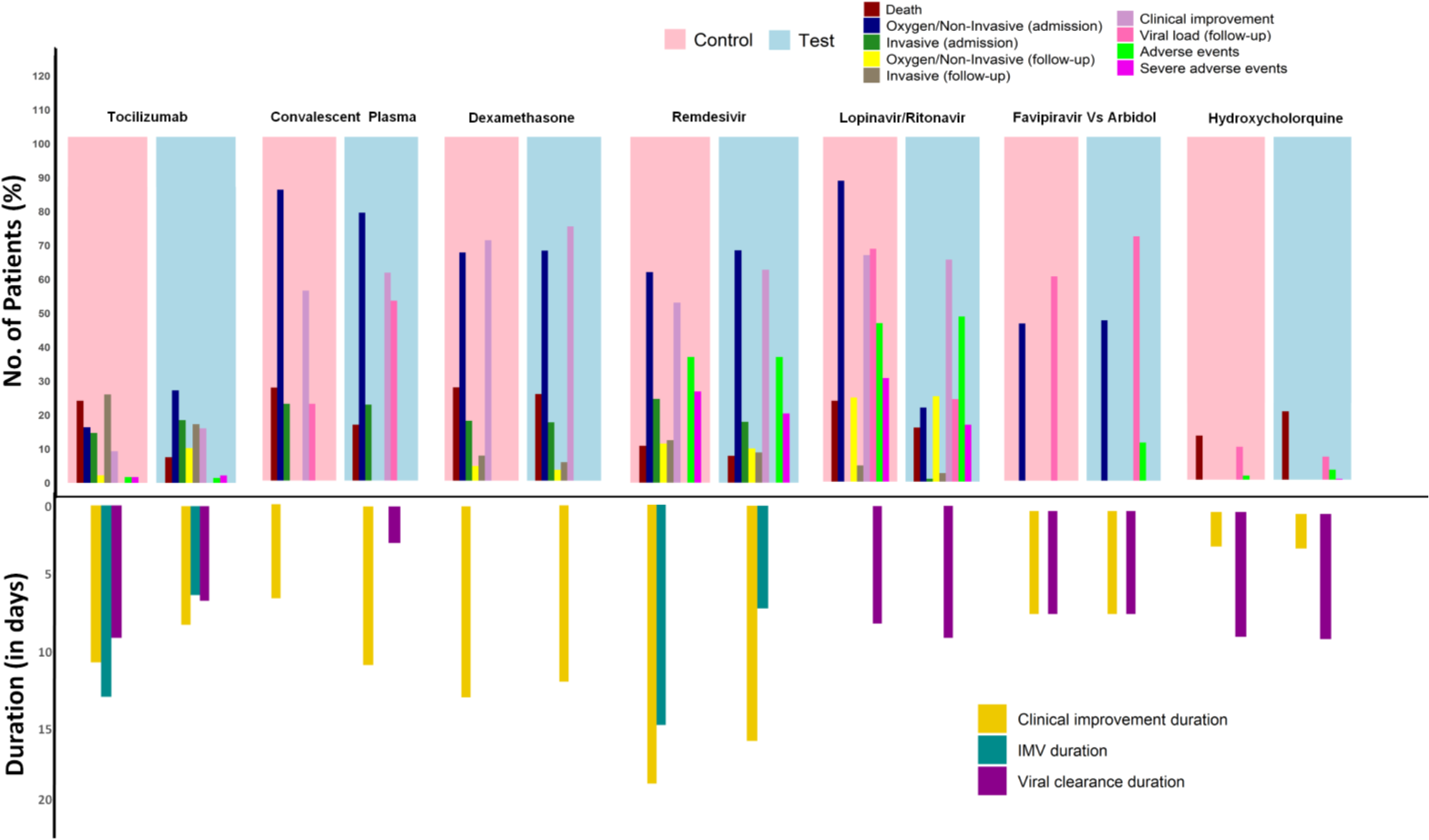
Histogram showing clinical outcomes of all the treatments comparing control and test groups. The top and bottom plot represents the percentage of patients and time period for each clinical outcome category respectively. The empty bars for few categories represent missing data

## 4. Discussion

Although mortality rate is 1% of COVID-19 patients [37], it has caused 991,224 deaths till now, and no therapeutic agent has received Food and Drug Administration (FDA) approval yet. There are many clinical studies reported on repurposed drugs for COVID-19 treatment. This meta-analysis indicates 29 of such studies involving 14,114 patients with COVID-19 to assess drug efficacy on the key outcomes, including mortality rate, viral clearance, clinical improvement, oxygen support, invasive ventilation, comorbid factors (diabetes, hypertension, heart disease, lung disease and renal disease) and adverse events.

According to the pooled results in subgroup analyses, assessing the severity of the disease, TCZ is found to be more effective in reducing the mortality rate (OR 27.50 [5.39-140.24]) of severe COVID-19 patients (Figure 1). TCZ, an approved antagonist of IL-6 receptor in rheumatoid arthritis with moderate to severe condition [38]. This is also considered as a selective cytokine inhibitor in COVID-19 [39], where multiple clinical studies have evaluated the safety and efficacy of TCZ in the severe stage of the disease.

In Capraa et al.’s study, a greater significance was seen in survival rate (p=0.004) and 92% recovery in TCZ treated patients as compared to the control [26]. But, Campochiaro’s study reveals no statistical difference in mortality rate (TCZ 15% *Vs*. control 33%) and clinical improvement (69% *Vs*. 61%), due to small cohort size [27]. In fact, two studies [35,40] concluded that early use and low dosage of TCZ has shown to be effective in reducing mortality rate, without any adverse events. In spite of that, three retrospective studies on TCZ [34,41,42], where 2 studied with cohort-control and 1 with single- arm, managed to show consistent results in relation to risk of IMV or death or ICU admission.

The first study has a larger cohort of 544 serious COVID-19 patients, showing a decrease in IMV risk up to 27% in the TCZ group than standard 41.5%. But, this study limits with a residual confounding that there were 4 patients with renal insufficiency or cancer in the TCZ group whereas it was 15 in the standard [41]. Similarly, the second study even with a smaller cohort could observe that TCZ treatment strongly reduces ICU admission or death (25% *Vs*. 72%, p = 0.002) or IMV support (0% *Vs*. 32%, P = 0.006) than standard care in the critically ill COVID-19 patients [42]. The third single-arm study with 100 patients also shows clinical improvement in 77% patients having respiratory support or 74% of 43 ICU patients by day-10 [34]. Additionally, two studies shows good improvement in the SpO2 levels (89% to 97%, day 1-10) [43] and PaO2:FiO2 ratio (152±53 to 302.2 ±126, day 0-14) of TCZ treatment [44].

A previous systematic review on TCZ reported 28 studies conducted in many different places; the results of most of these studies were favorable to TCZ therapy in severe and critically ill COVID-19 patients [45]. Based on this, TCZ is safe in treating both early and late stage respiratory conditions of COVID-19, but so far, no RCT has been reported in treating the severe groups, where the RCT is still on-going to measure the efficacy and safety of this treatment in case of severe COVID-19 [46].

CP is another type of immune therapy that shows less or no significance (OR 2.12 [0.70-6.38]; 1.70 [0.63-4.59]) in the subgrouping of 2 RCTs included. Results from a RCT using CP have shown that mortality rate (p=0.95) and improvement of day-15 disease severity (p=0.58) were not significant when compared to the standard group. The study also shows that COVID-19 patients already have enough neutralizing antibody titers than donors and suggests the use of CP at early symptom onset [28]. Another RCT also showed similar results, besides, they showed a higher significance (87.2% CP *Vs*. 37.5% standard) in viral load negativity within 72 hrs after CP transfusion in severe COVID-19 patients [29]. Limitation in both studies was that they were soon terminated before attaining enough sample size which prevented definite conclusions of clinical benefits.

In our post-search, a large RCT done on CP management was found. This enrolled almost 464 moderately ill COVID-19 patients, and showed results in line with 2 previous RCTs, that mortality was not significantly different between CP treated and control groups 14.5% *Vs*. 13.5% [47]. According to these reports, CP therapy has shown to be more efficient in early negative conversion of viral RNA, but no effect in reducing the mortality rate of moderate-severe COVID-19 patients.

RM shows less or no significance in our subgroup analyses (OR 1.81 [1.15-2.83]; 0.91 [0.41- 2.03]. This shows that it is not effective in reducing the mortality rate for severe COVID-19 patients. Two RCTs with placebo-controlled have been reported, one [30] of which has a larger study size with improvised protocol than the other [31]. The first study [30] favors RM (a 10-day course) over placebo with short recovery time (11 *Vs*. 15 days) for all clinical outcomes except patients on IMV or ECMO. They further suggest that it is supportive for hospitalized and low supplemental oxygen support patients. A second study [31] trialed 2:1 randomization with limited sample shows no significant reduction of viral load or duration of clinical improvement in severe COVID-19 patients. Both RCTs describe that the mortality rates were insignificant between treatment and placebo. Also recommend that RM could be given as an antiviral agent in combination with other therapy before the condition progresses to need for IMV [30,31].

Subsequently, Gein et al. study on compassionate-use of RM, shows 22% death and 70% clinical improvement among patients. Also, 50% (25/53) of severe patients with age group 70 or older were discharged. However, adverse events occurred in 60% of patients (such as elevated hepatic enzymes, rash, hypotension and renal impairment) and serious adverse events in 23% (multiple-organ-dysfunction syndrome, acute kidney injury, septic shock and hypotension), which were caused by RM treatment alone [48,49].

Two other studies [50,51], randomized moderate and severe patients with RM for either 5-day or 10-day courses. The clinical improvement of severe patients (without receiving mechanical ventilation) was not significant (64% *Vs*. 54%) between the two courses [50]. Anyhow, in patients with moderate severity, the 5-day course had statistically significant clinical status when compared to standard care (OR 1.65 95% CI, 1.09-2.48; p = .02), whereas the 10-day course showed no effect [51]. Majority of these study outcomes suggest that RM can be used for hospitalized and lower respiratory tract infectious COVID-19 patients.

Regarding DM, Horby et al. have published a well-designed largest RCT that enrolled the highest population size (n=6425), when compared to any other studies included in this meta-analysis. The study shows a low dose of DM is most effective in patients on IMV or ECMO than without respiratory support, where the risk of death is reduced significantly in DM group than usual care (29.3% *Vs*. 41.4%) of critically ill COVID-19 patients. However, the overall mortality rate of all groups including those without respiratory support does not seem to have much difference (22.9% *Vs*. 25.7%) [32], which is why the subgroup analysis in our study shows a marginal effect in mortality rate (OR 1.19 [1.05-1.35]). Therefore, DM could be recommended in treating long term symptom COVID-19 patients, requiring IMV than recent symptom onset.

The studies on HCQ and antivirals (LPV-r, arbidol, FPV) in mild-moderate subgroups show a null effect (OR 1.10 [0.29-4.11]) (Figure 1), since no event of death occurred in both control and treatment groups. However, many other clinical outcomes related to these drugs are reported by the trials. HCQ, an anti-malarial agent presumed to exert antiviral and immunomodulating effects in COVID-19. But, Geleris et al. studied a large cohort (n=1376) of hospitalized COVID- 19 patients, suggested that HCQ use had no potency for end stage intubation or death (hazard ratio, 1.04; 95% CI, 0.82 to 1.32) [33]. This study had a poor score (OR 0.64 [0.47-0.86]) in the severe subgroup (Figure 1). Two RCTs from China studied HCQ with a small number of mild-moderate patients. The first is a preprint [52] and only partially confirmed the drug effect on symptomatic outcome (like fever and cough). The other study [53] shows no significance in viral clearance between HCQ plus standard *Vs*. standard alone treatment, in addition, more adverse events were found high in HCQ group. Recent research shows HCQ inhibits trained immunity by an epigenetic modulation. This prevents the antiviral effects of the bodily innate immune response against the SARS-CoV-2 infection [54]. Together, these results indicate that HCQ is not a promising rescue option for mild-moderate and severe COVID-19 patients.

LPV-r or Arbidol alone shows little benefit for clinical outcome of mild-moderate COVID-19 patients [55], but Deng’s study showed that combined effect is more favorable [56]. A retrospective study commends, LPV-r has a more rapid effect in viral clearance than HCQ [57]. Although Cao et al. performed a RCT on LPV-r found no benefit in mortality reduction or receiving oxygen support in case of severe COVID-19 patients. Thus no effect was seen in the mortality rate (OR 1.40 [0.71-2.76]) of the severe subgroup (Figure 1). Their finding says the positive viral RNA appeared till the end of trial in LPV-r group (40.7%), but it was not confirmed with the presence of viral infection. The study limits without blinding which might have influenced the outcome and premature discontinuation (14%) of the treatment [58]. Therefore, the LPV-r treatment seems to have antiviral benefits during mild COVID-19 condition but has no benefit in mortality reduction.

FPV is an inhibitor of RNA-dependent RNA polymerase of viral RNA, evaluated by in-vitro studies to be active against COVID-19. An observational study with 2158 COVID-19 patients, studied FPV use on compassionate basis. They show mortality was higher in the severe group (31.7%) when compared to mild (5.1%) and moderate (12.7%) by day-30. Likewise, the rate of recovery was lower in the severe group (14.7%) than the other two conditions (61.7% and 42.7%). Also, deaths were frequent in elderly and 24.65% of patients posed adverse events with FPV therapy. The study unfortunately is limited since it contains no background data of patients though they are registered from 407 hospitals [59]. A study in Thailand [36] favors the use of FPV with 100% of clinical improvement in 27 hospitalized patients (without oxygen support) and 83.3% in 30 serious conditions (requiring supplemental oxygen or IMV) at day 28. However, the study is limited by the use of additional agents such as chloroquine and hence could not find the actual impact of FPV. Another study [60] shows, by day-7, FPV had no difference in recovery rate when compared with Arbidol therapy for moderate COVID-19 patients.

Synthetic serine protease inhibitors (serpins) were earlier used to treat diseases like Acute myocardial infarction, Ischemic stroke and Pulmonary embolism [61]. More recently Nafamostat, a Serpin was found to prevent the proteolytic activity of transmembrane protease serine 2 (TMPRSS2) and thus inhibits the viral fusion with the host cells [62,63]. The combination of two therapies (serpin and FPV), may allow the inhibition of viral entry as well as the replication. A small case series shows that FPV in combination with Nafamostat was found to have benefits on severe COVID-19. In which, 8 of 11 patients were extubated and 9 discharged from ICU. But still there is no convincing evidence supporting this study [64]. Taken together, FPV use is efficacious in mild-moderate COVID-19 patients. But for severe or critically ill conditions, the addition of Serpins with the FPV may be helpful.

In summary i) TCZ is effective in severe COVID-19, but requires a RCT to validate the results. ii) DM works only on patients with IMV in reducing mortality, however several unresolved questions still exist. iii) RM benefits patients without having IMV support with shorter recovery time. iv) CP and LPV-r can cure only mild illness and CP was found to be efficacious in bringing down the viral load in at least hours. Both have no benefit in mortality reduction v) HCQ has no potency or antiviral effects. vi) FPV when used in combination with serpins found to have additive effects on severe COVID-19, but more findings are required to assess the drug tolerability.

One previous meta-analysis [65] was reported already on repurposed drugs by June 9 (pre-print), however the current study has added more therapeutic agents with its recent trials including a huge number of patients. Our study is bound by limitations of much high risk quality evidence. In addition, a few non-comparable single-arm studies and smaller cohorts are underpowered to assess the clinical outcomes that were addressed.

## 5. Conclusion

Based on the available evidence, this analysis has found immunotherapy (TCZ) was superior to antivirals in most of the clinical outcomes. TCZ has promising effect in both early and late stages of disease severity. Although, the RCT with TCZ and other monoclonal antibody cocktails in the 6-arm RECOVERY clinical trial (NCT04381936) are still on-going, the results of which may provide an optimal dosage and safety details for an appropriate benefit of the therapy. The meta-regression analysis shows TCZ exposure in patients with hypertension comorbid condition had a significant correlation with risk of non-invasive oxygen support (P=0.02). On the other hand, lung diseases (comorbidity) had lower chance of requiring non-invasive support. A low dose corticosteroid (DM) had only a marginal effect on overall motarlity rate. However, based on the evidence, it is a good rescue option for patients on IMV. HCQ was found to have no clinical effects for COVID-19, whereas it plays a role in inhibiting the trained immune system. LPV-r, not efficacious in severe COVID-19, yet has some antiviral effects in mild conditions. The use of CP rapidly reduces viral load within a few hours in all stages of COVID-19. The results of HCQ and LPV-r were in line with the partially reported recent 6-arm trial (NCT04381936). The Antivirals RM and FPV were recommended as a combinatorial therapy with other agents. A 10- day course of RM has speedy recovery in hospitalized patients (without IMV) and a 5-day course is safe in moderate COVID-19. FPV alone treats patients before the end stage of COVID-19 patients. But the combinatorial use of Serpins with FPV may be efficacious for even critical patients. Furthermore, high quality evidence is required to evaluate its usage in severe or critical COVID-19.

## Supporting information

PRISMA 2009 checklist

## Data Availability

Data can be accessed on request

## Acknowledgment

We thank Abhimanyu Swaroop, IIT-Madras for assistance with screening of articles during the initial phase of study.

## Authors’ contributions

T.T and G.S devised the study. E.J.E, A.K and R.M searched the database, extracted the data. E.J.E and V.S.K reviewed and planned the analysis for included articles. V.S.K performed the statistical analysis and designed the figures. E.J.E and V.S.K wrote the initial draft. T.T and G.S revised and edited the paper.

## Conflict of interest

The authors declare that they have no competing financial interests or personal relationships that could have appeared to influence the work reported in this paper.

## Funding

There was no funding provided for this work

## Notes

### Competing Interest Statement

The authors have declared no competing interest.

### Funding Statement

The authors received no funding for this work.

### Author Declarations

This analysis do not require approval from ethical guidelines as the work did not involve patients directly

