## Supplemental Table 1 for "Rationale and prognosis of repurposed drugs with risk stratification of COVID-19 patients requiring Oxygen supplementation: A systematic review and meta-analysis"

**Supplemental information**

**Table S1. Risk of bias assessment of 8 single-arm studies with Newcastle–Ottawa scale**

| **Author** | **Representativeness** | **Selection of the non-exposed cohort** | **Ascertainment of exposure** | **Demonstration** | **Comparability** | **Assessment of outcome** | **Follow-up** | **Adequacy of follow-up** | **Total score** |
| --- | --- | --- | --- | --- | --- | --- | --- | --- | --- |
| Grein | C | NA | A* | A* | C* | A* | A* | A* | 6 |
| Rattanaumpawan | C | NA | A* | A* | C* | A* | A* | A* | 6 |
| Doi | C | NA | C | A* | C* | A* | A* | B* | 5 |
| Doi | C | NA | D | B | C* | D | A* | A* | 3 |
| Tomasiewicz | C | NA | C | A* | C* | A* | A* | A* | 5 |
| Sciascia | C | NA | D | A* | C* | A* | A* | A* | 5 |
| Toniati | C | NA | C | A* | C* | D | B | A* | 3 |
| Montalva | C | NA | A* | A* | C* | A* | B | D | 4 |
